## Supplementary figures and images for "Localized skin inflammation during cutaneous leishmaniasis drives a chronic, systemic IFN-γ signature"

### Supplemental Figure 1

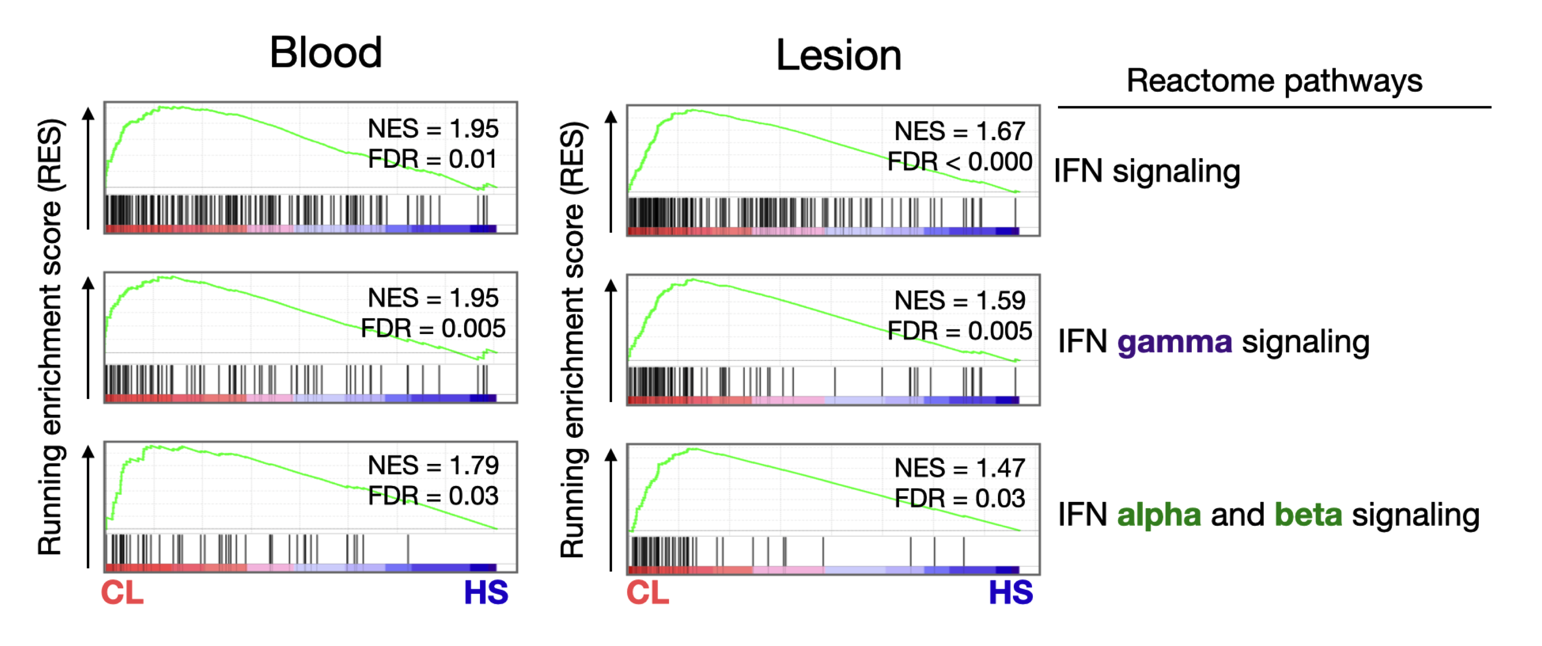

### Supplemental Figure 2

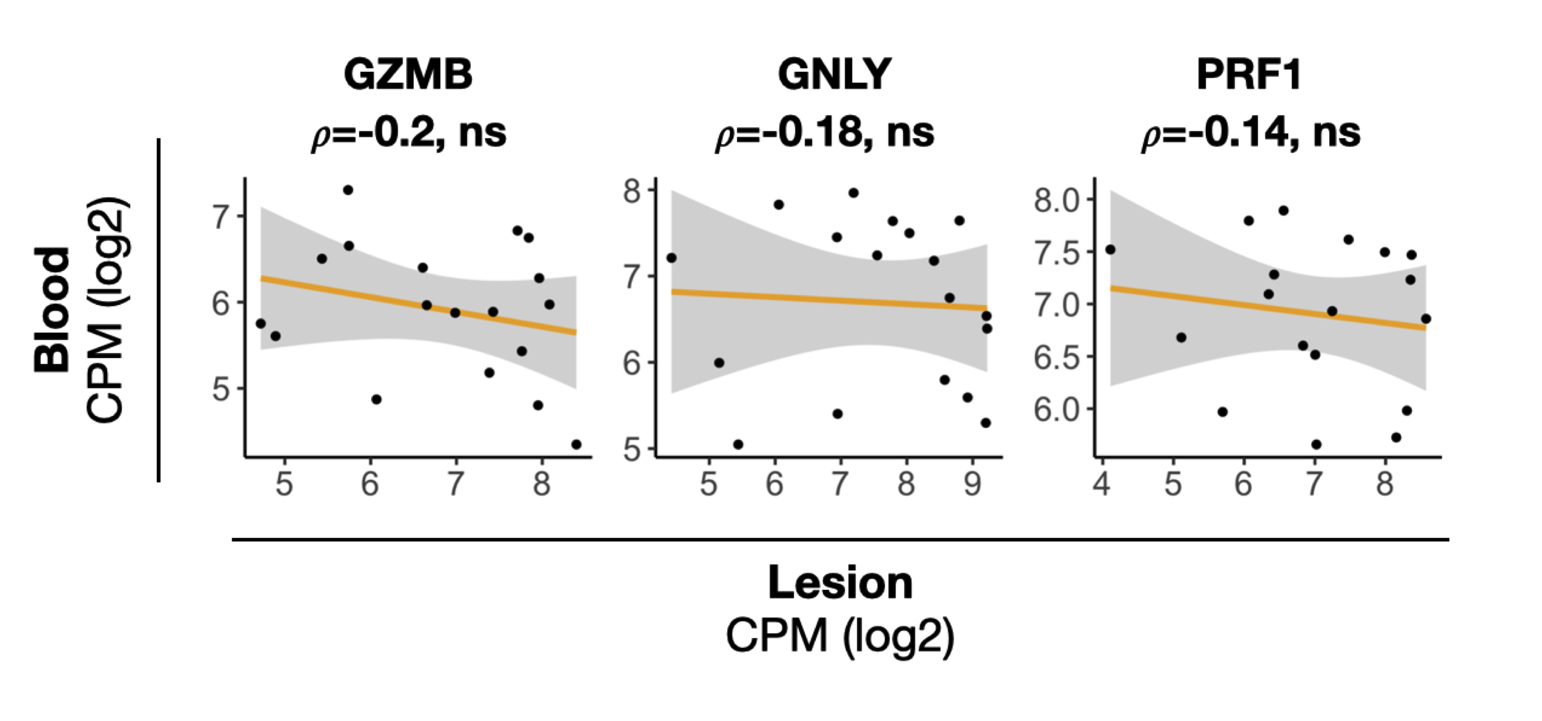

### Supplemental Figure 3

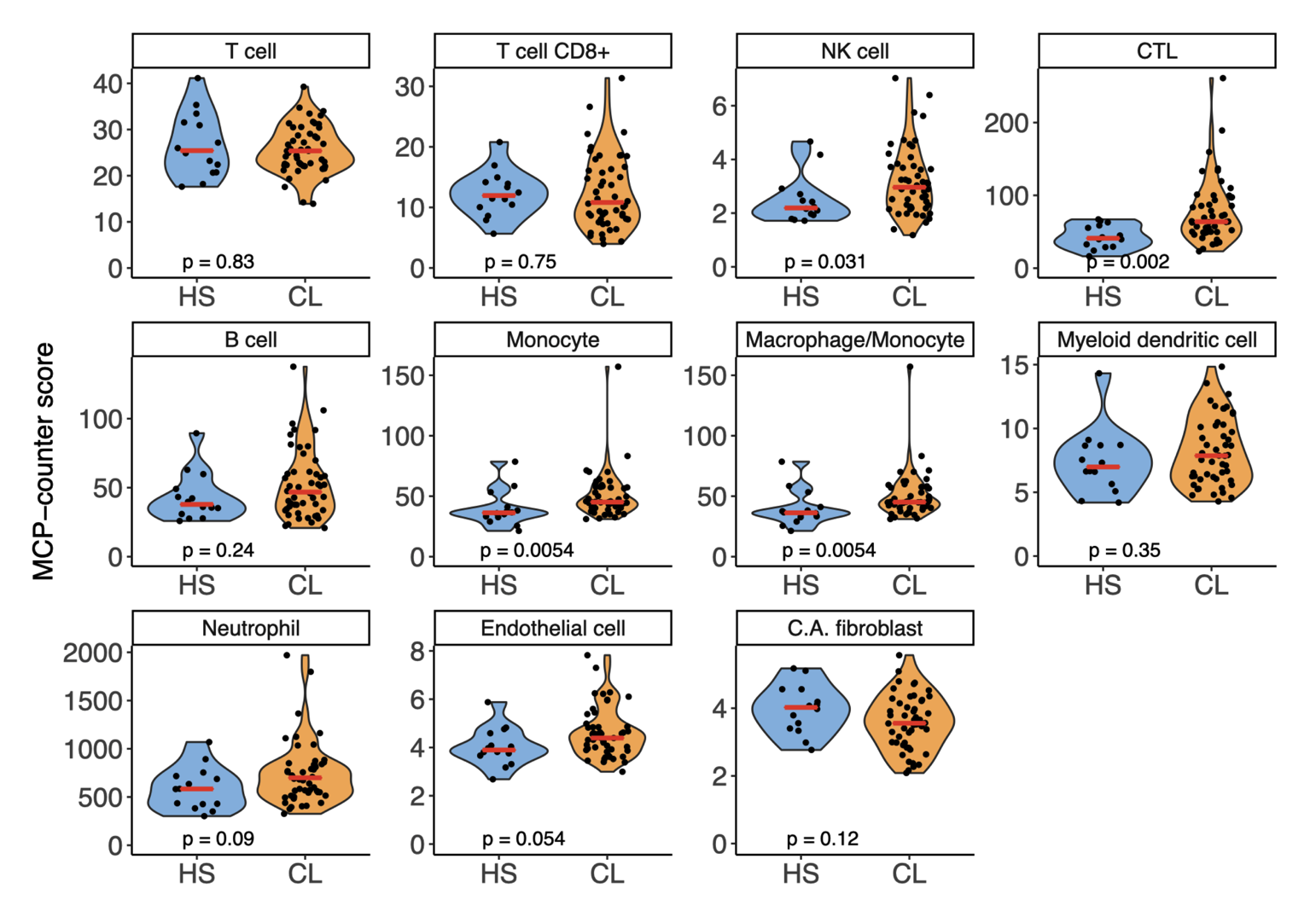

### Supplemental Figure 4

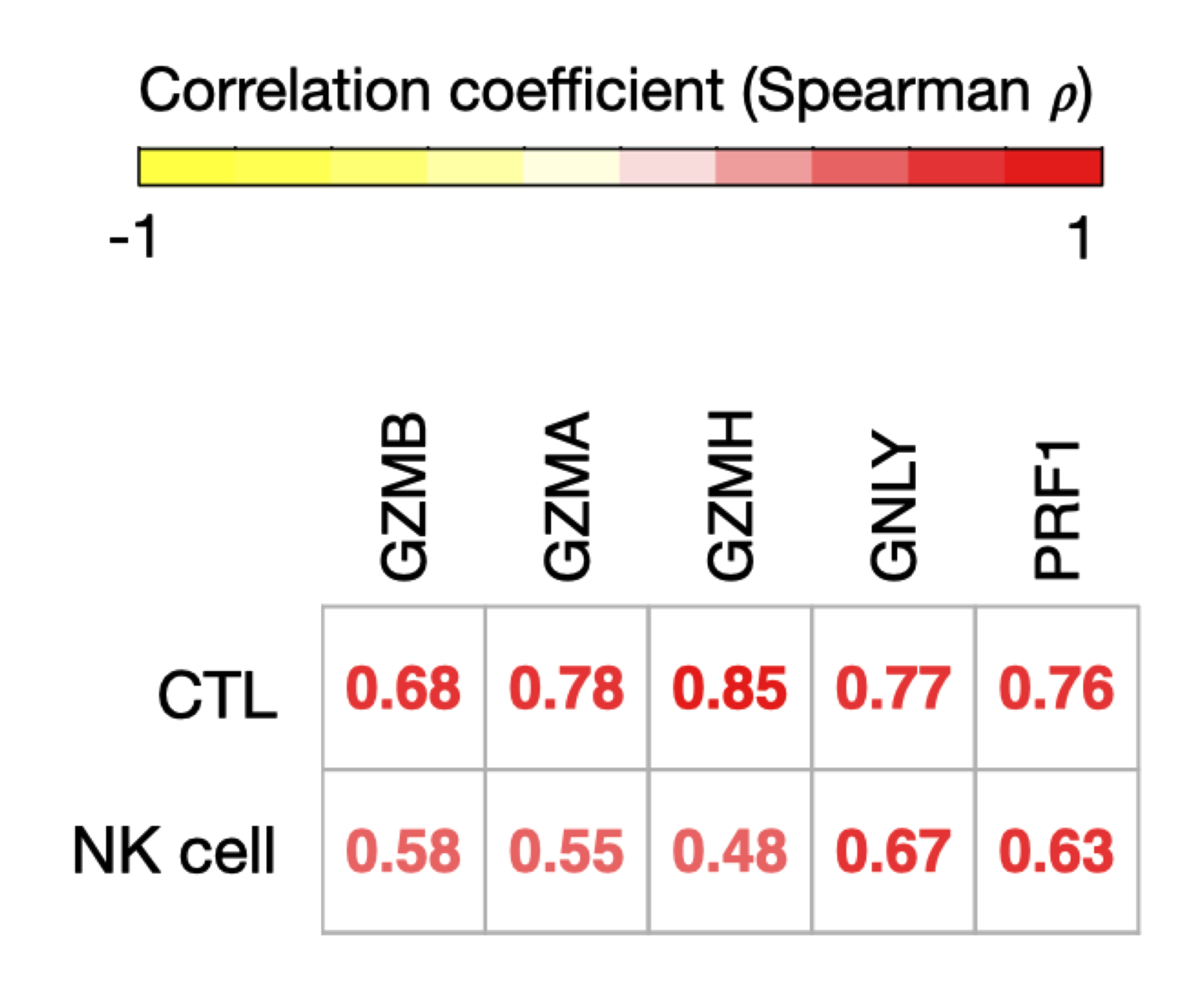

### Supplemental Figure 5

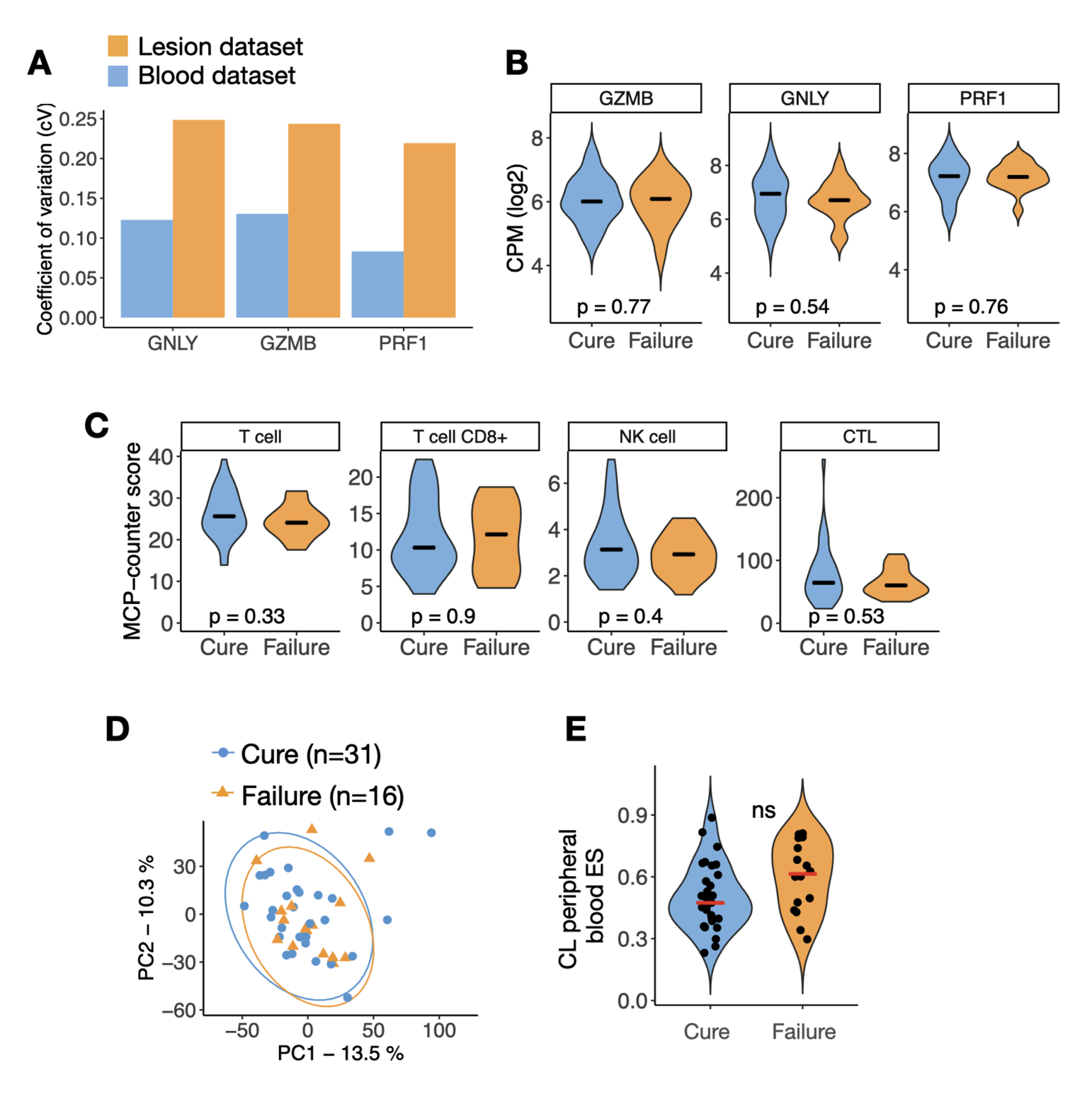

### Supplemental Figure 6

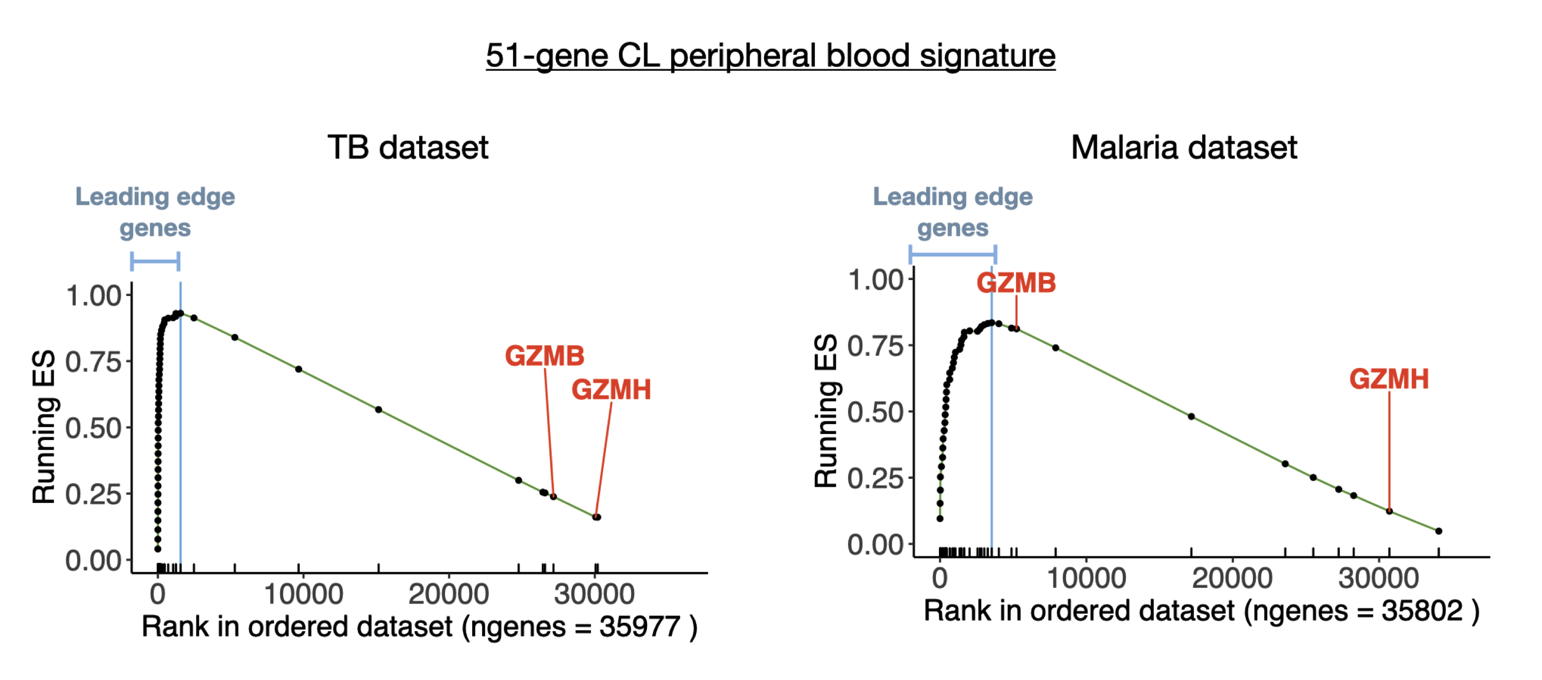
